# Strengthening support for cancer caregivers: an investigation of caregiving perceptions and informational needs

**DOI:** 10.1101/2025.07.18.25331798

**Authors:** Diana C. Dima, Meredith E. Giuliani, Tulin D. Cil, Jennifer Deering, Jennifer Jones, Andrew Matthew, Rinat Nissim, Tina Papadakos, Naa Kwarley Quartey, Gary Rodin, Andrew Stechkevich, Santhosh Thyagu, Mohamed Ugas, Yuan Zhong, Janet Papadakos

## Abstract

**Introduction:** Unpaid caregivers play a key role in supporting people with cancer. However, their needs and perspectives are often overlooked, and few healthcare institutions offer dedicated support programs. To address this, we assessed and compared the perceptions and informational needs of unpaid caregivers and cancer patients.

**Methods:** Caregivers (N=115) and cancer patients (N=99) from a large urban cancer centre completed a one-time survey evaluating their perceptions of the medical care and caregiver support they had received, their informational needs, as well as their health literacy and computer proficiency. Caregivers also completed validated scales evaluating caregiver burden, preparedness, competence, and reward.

We compared patients’ and caregivers’ reported perceptions and analyzed their informational needs across knowledge domains. We conducted stepwise regression to understand the relationship between participant characteristics and scores, including health literacy and caregiving burden, and their informational needs.

**Results:** Patients underestimated many aspects of caregivers’ experience, and caregivers were less satisfied than patients with the medical care received. Caregivers with higher health literacy had lower informational needs and caregiver burden. Both groups expressed a strong need for medical information in addition to psychological, social, and practical support.

**Conclusion:** These results reveal a pressing need for proactive medical and practical training for caregivers to complement psychological interventions. Furthermore, interventions to improve public health literacy may enable those who become caregivers to better navigate the challenges of their role.

## 1. Introduction

Caregivers providing unpaid care for a family member or friend play an increasingly important part in healthcare. Cancer often requires unpaid care^1^, and cancer caregivers take on a wide range of tasks, from communicating health information and supporting patients with daily activities, to managing medication and symptoms^2^. Due to the increasing shift of cancer care to outpatient settings, these tasks are increasingly complex and often require clinical knowledge and skill^3^. Yet support programs and resources for caregivers can be challenging to implement, given the large variations in caregivers’ experience and the paucity of data. Existing interventions focus almost exclusively on psychological outcomes and often require synchronous attendance, which can reduce accessibility^4,5^.

Caregiving is a complex experience characterized by both rewards and challenges. The impacts of caregiving depend on socio-economic, cultural, and lifestyle factors, and can include damaging effects on caregivers’ financial, social, physical, and mental health^6,7^. The cancer journey involves unique challenges, such as sudden deteriorations in health, concerns about disease recurrence, and changes in caregivers’ role over time^8,9^. Particularly in intermediate and advanced cancer phases, cancer caregivers suffer from many unmet needs, and their distress often remains unaddressed^10^. They often feel unprepared and isolated in their new role, and need specific knowledge to face medical, practical, social, and psychological challenges^11–13^. These needs are amplified by their unique status as “information brokers” who manage the flow of information between healthcare providers, the patient, and the patient’s social circle^14^.

Caregivers’ informational needs are different from those of patients, yet many caregiver programs remain limited in scope and accessibility^15,16^. Furthermore, caregivers and patients report diverging perceptions: while caregivers may overestimate the patients’ distress and pain, patients may misjudge caregivers’ burden^17,18^. These differences in perception can heighten caregivers’ stress and make communication with patients more difficult.

We conducted a cross-sectional assessment of caregiving-related perceptions in cancer caregivers and patients in order to investigate differences in their perceptions of caregiving, caregiver support, and medical care. We also evaluated caregivers’ and patients’ informational needs across the medical, practical, physical, social, psychological, and spiritual domains. Finally, we identified the factors associated with caregivers’s experience and informational needs. This rich dataset highlights the importance of health literacy and proactive educational interventions in supporting cancer caregivers.

## 2. Methods

### 2.1. Sampling and recruitment

Cancer patients and caregivers visiting five outpatient clinics (Lung, Hematology, Head and Neck, Gynecology, and Gastrointestinal clinics) at the Princess Margaret Cancer Centre in Toronto, Ontario, Canada were invited to complete a cross-sectional survey between July 2021 and May 2022. Patients and caregivers were considered eligible if they were at least 18, spoke English, and had a confirmed cancer diagnosis or were caring for someone with a cancer diagnosis. Paper questionnaires were completed in clinic, with minimal supervision from the study team, or at home, and returned to the clinic or the Patient and Family Library at the hospital. Questionnaires were also available electronically, but no participants chose this method. Among participants who agreed to take the survey, the proportion of completed surveys was 43% (patients) and 45% (caregivers). Consent was implied by completion of the questionnaire, and participation was voluntary, optional, and anonymous. Approval to conduct the study was obtained from the Institutional Research Ethics Board (REB#: 09-0952).

### 2.2. Survey design

The survey design was informed by an in-depth literature review and consultation with patient educators and clinical subject matter experts. To promote readability, all questions were written in plain language^19^. Survey items (except validated scales) were tested for face validity with five patients, ensuring that questions were understood as intended. Two versions of the survey were distributed to patients and caregivers (see **Supplementary Information**). The versions differed in wording, ensuring that patients evaluated caregivers’ informational needs, and contained additional validated scales for caregivers. The surveys consisted of the following sections:

1. Demographic and health information (14 items). This section collected demographic information about participants’ age, gender, place of birth, ethnicity, English language fluency, education, employment status, household income, and Internet access. Health information included cancer type, cancer stage, and purpose of treatment. Caregivers were also asked whether they were a main caregiver, while patients were asked if they had someone to help manage their care. Both groups also estimated the time they/their family member spent on caregiving and transportation to appointments, as well as the amount of help they/their family member needed with daily activities.
2. Measures of health literacy (16 items) and computer proficiency (12 items). Participants completed the validated European Health Literacy Survey Questionnaire (HLS-EU-Q16)^20^ and the validated Computer Proficiency Questionnaire (CPQ-12)^21^.
3. Validated caregiving scales. Caregivers completed the Burden Scale for Family Caregivers (BSFC)^22^, the Preparedness for Caregiving Scale (PCS), the Caregiver Competence Scale (CCS), and the Rewards of Caregiving Scale (RCS)^23^.
4. The impact of cancer on caregivers (7 items). Caregivers and patients evaluated the extent to which their loved one’s cancer affected caregivers in terms of stress, physical health, time for family and friends, as well as attitudes and values. Questions were answered on a three-point scale (“No, not at all,” “Yes, a little”, and “Yes, a lot”), and included a “Do not know/Not relevant” option.
5. Satisfaction with medical care received (20/13 items). Caregivers and patients rated their satisfaction on 20 and 13 items respectively, each related to the medical care received. Items included pain relief, referrals, coordination of care, information received, and the availability of healthcare professionals.
6. Informational needs (67 items). Caregivers and patients rated their informational needs across six domains of need adapted from Steele and Fitch^24^: medical (e.g. treatment options, health information); practical (e.g. transportation, legal and financial support); physical (e.g. pain, symptoms, side effects); social (e.g. relationships, communicating with others); psychological (e.g. coping with grief, uncertainty, changes in body image); and spiritual (e.g. the value of spirituality and spiritual services). Participants rated each item on a Likert scale from 1 (“Not important”) to 3 (“Very important”) or selected “Not applicable”.
7. Information delivery preferences (6 items). For each knowledge domain, participants selected their top three choices of information delivery method from a list including one-to-one teaching, group classes, support groups, online forums, online video and audio materials, DVDs, books, and pamphlets.
8. Satisfaction with the caregiver support received (6 items). Participants reported how often hospital healthcare workers showed an interest in how caregivers felt, involved them in their family member’s care, and offered help. They also rated the amount of time spent giving caregiving information, whether the information was provided in a sensitive way, and how well the hospital had informed caregivers overall. Questions were answered on different three- or four-point scales.
9. Open-ended questions. Questions with comment-style answers enabled participants to elaborate on their highest informational need, their satisfaction with the information they had received, their preferred modality of information delivery, and any additional comments or suggestions.

### 2.3. Statistical analysis

Demographic and health information was summarized using descriptive statistics. Informational needs data were used to calculate importance scores for each domain, defined as the percentage of items rated as “Very important”. Participants’ satisfaction with the medical care received was summarized using a similar satisfaction measure, consisting of the percentage of items with which participants were “Very satisfied.” Modalities of information delivery were ranked according to the proportion of times they were selected as a top choice.

Associations between participant characteristics, scores on validated scales (health literacy, computer proficiency, and caregiving scales), and informational importance scores were analyzed using pairwise Pearson’s correlations. Next, a multivariate regression analysis was conducted for each knowledge domain, with the importance scores as response variables. Predictors included participant characteristics (age, gender, place of birth, English level, education level, main activity, income, cancer type and phase, treatment purpose), caregiving survey responses (time spent caregiving/transporting, amount of help needed by patient) and scores on validated scales (health literacy, computer proficiency, and caregiving scales). We used a stepwise variable selection method to iteratively add or remove predictors from the full regression model so as to minimize the Bayesian Information Criterion (BIC), which balances model fit and complexity.

Open-ended responses were analyzed using a qualitative coding approach. Responses were carefully reviewed and thematically categorized based on recurring patterns. Key responses were selected and reported to highlight significant findings.

## 3. Results

### 3.1. Health literacy and computer proficiency

The survey was completed by 115 caregivers (74 female) and 99 patients (50 female; see **Table 1** for demographic and health information). On average, participants had sufficient health literacy as measured on the HLS-EU-Q-16 scale (over 12; **Supplementary Figure 1**). Scores did not significantly differ between caregivers (12.8±3.3) and patients (12.1±4.1; t(154.7)=1.25, *p*=0.21). Computer proficiency scores were high on average in both samples (28.4±3 and 24.5±6.8 respectively with a maximum of 30) and were significantly higher in caregivers than patients (t(111.9) = 4.98, *p*<0.001).

**Table 1.** Demographic and health information. Caregivers: N = 115; Patients: N = 99.

| Variable |  | Caregivers |  | Patients |  |
| --- | --- | --- | --- | --- | --- |
| Age | Median ±SD | 55±14.9 | | 67.5±14.6 | |
|  | Range | 13-83 |  | 19-89 |  |
|  | Category | Num | % | Num | % |
| Gender | Women | 74 | 65 | 50 | 51 |
|  | Men | 40 | 35 | 49 | 49 |
| Place of birth | Canada | 79 | 69 | 51 | 53 |
|  | Other | 36 | 31 | 46 | 47 |
| Ethnicity | Indigenous | 1 | 1 | 4 | 4 |
|  | Arab/West Asian | 1 | 1 | 0 | 0 |
|  | Black/African | 2 | 2 | 3 | 3 |
|  | East Asian | 12 | 10 | 9 | 9 |
|  | Latin American/Latino | 1 | 1 | 3 | 3 |
|  | South Asian | 5 | 4 | 2 | 2 |
|  | Southeast Asian | 4 | 4 | 5 | 5 |
|  | White/Caucasian/European | 84 | 74 | 68 | 69 |
|  | Other | 4 | 3 | 4 | 4 |
| English level | Not fluent | 1 | 1 | 14 | 14 |
|  | Fluent | 20 | 17 | 24 | 24 |
|  | Very fluent | 94 | 82 | 61 | 62 |
| Comfortable receiving health information in English | Yes | 115 | 100 | 89 | 92 |
|  | No | 0 | 0 | 8 | 8 |
| Comfortable filling out medical forms in English | Yes | 114 | 99 | 79 | 83 |
|  | No | 1 | 1 | 8 | 8 |
| Education level | High school or less | 15 | 13 | 40 | 40 |
|  | Some college/university | 14 | 12 | 9 | 10 |
|  | College/university | 55 | 48 | 35 | 35 |
|  | Graduate school | 26 | 23 | 15 | 15 |
|  | Other | 5 | 4 | 0 | 0 |
| Main activity | Working | 64 | 56 | 50 | 51 |
|  | Retired | 37 | 32 | 35 | 36 |
|  | Other | 14 | 12 | 13 | 13 |
| Household income | < \$49,999 | 11 | 10 | 24 | 25 |
| | \$50,000-99,999 | 37 | 33 | 23 | 24 |
| | >\$99,999 | 44 | 39 | 29 | 30 |
|  | Prefer not to answer | 21 | 19 | 20 | 21 |
| Have easy access to the Internet to look up health information | Yes | 114 | 99 | 97 | 99 |
|  | No | 1 | 1 | 1 | 1 |
| Type of cancer | Breast | 3 | 3 | 2 | 2 |
|  | Gastrointestinal | 32 | 29 | 28 | 29 |
|  | Genitourinary | 1 | 1 | 2 | 2 |
|  | Gynecological | 14 | 12 | 12 | 12 |
|  | Head and Neck | 14 | 13 | 12 | 12 |
|  | Hematologic | 25 | 22 | 21 | 22 |
|  | Other | 22 | 20 | 20 | 21 |
| Point in cancer journey | Newly diagnosed | 13 | 12 | 12 | 13 |
|  | Ongoing treatment | 61 | 54 | 47 | 51 |
|  | Follow-up care | 39 | 35 | 34 | 36 |
| Cancer treatment purpose | Palliative | 11 | 9 | 8 | 8 |
|  | To cure cancer | 52 | 41 | 46 | 47 |
|  | To prolong survival | 52 | 42 | 40 | 41 |
|  | Do not know | 10 | 8 | 4 | 4 |

### 3.2. Perceptions of time spent caregiving

In both samples, 51% of participants estimated caregivers’ workload at over 15 hours per week, and over 75% reported that caregivers spent “a lot of time” transporting patients to appointments. When asked how much help the patient needed with daily activities, perceptions were different: 57% of patients reported that no help was needed, while 39% of caregivers reported that several hours each week were spent caregiving (**Figure 1a**). Note that these are group-level differences, as we did not require or record the presence of patient-caregiver dyads in our data.

**Figure 1.**
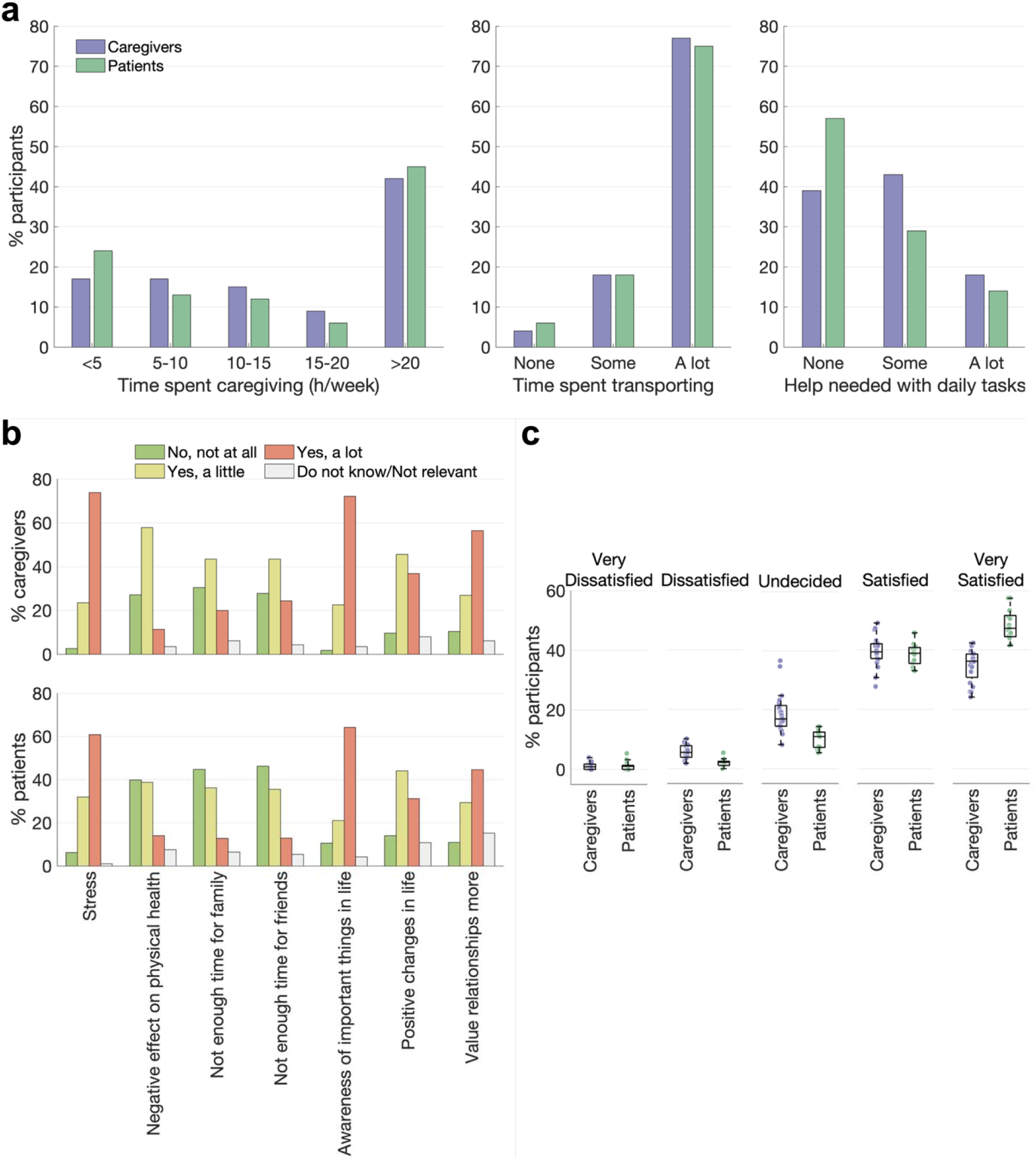
Caregiving perceptions among survey respondents. **a.** Perceived time spent caregiving. Eighty-eight percent of caregivers surveyed (n=101) reported being the main caregiver for their family member or friend, while 98% of patients (n=97) reported having someone who helps them manage their care. **b.** Perceived impact of cancer on caregivers. Bars show the percentage of participants choosing each answer option. **c.** Satisfaction with the medical care received by the patient. Individual dots are items in the satisfaction portion of the survey. Boxplots display the median proportion of participants choosing each answer across all questions.

### 3.5. The impact of cancer on caregivers

Most caregivers (74%) reported that their family member or friend’s cancer caused them a lot of stress, as well as made them aware of the important things in life (72%). Many patients also noted these impacts on caregivers (61% and 64% respectively). However, patients underestimated all impacts of cancer as reported by caregivers, including negative impacts on physical health and time (**Figure 1b**).

### 3.6. Satisfaction with the medical care received

Overall, most participants reported being “Satisfied” or “Very satisfied” with the healthcare patients had received (**Figure 1c**). However, satisfaction scores, computed as the percentage of items with which responders were “Very satisfied”, were significantly higher for patients (47.6±41.5) than for caregivers (34.8±36.1; *t (*189.8) =2.35, CI [2.1, 23.5], *p*=0.02).

### 3.7. Satisfaction with the caregiver support received

Most caregivers and patients (over 70%) reported that caregivers were well informed by the hospital (**Supplementary Table 1**). However, when asked if healthcare professionals had noticed when caregivers were not doing well and offered help, 30.8% of caregivers and 24.1% of patients thought this happened “rarely/never”. When asked if healthcare workers had shown an interest in how caregivers were feeling, 32.7% of caregivers and 26.7% of patients responded “rarely/never”.

### 3.4. Caregiving measures

Caregivers reported medium levels of caregiver burden (BSFC: 13.7±6, maximum 30) and preparedness (PCS: 17.4±8.4, maximum 32) and relatively high levels of perceived competence (CCS: 7.8±2.7, maximum 12) and reward (RCS: 28.4±8.5, maximum 40).

Women reported being more burdened by caregiving than men (**Figure 2**; t (70.7) = 3.14, CI [1.4, 6.1], *p*=0.0025). This was also reflected in a higher proportion of women reporting they spent over 15 hours per week caregiving (54% of women compared to 40% of men).

**Figure 2.**
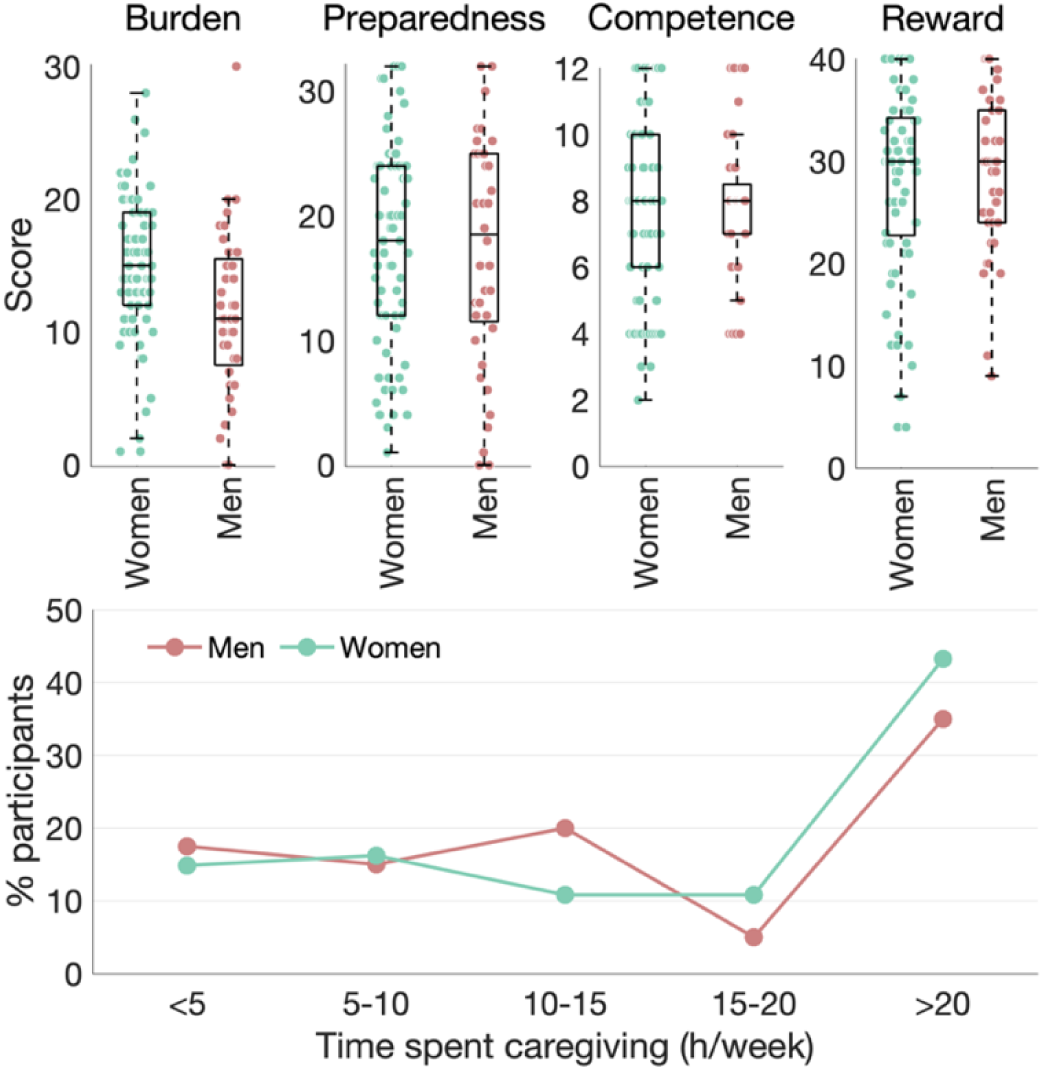
Gender differences in caregiving. **Top:** Participant scores on four validated caregiving scales, split by gender. Dots are individual scores, while boxplots show median scores. **Bottom:** Gender differences in self-reported time spent caregiving. Dots show the proportions of men and women who selected each response.

### 3.8. Informational needs

We assessed caregiving-related informational needs across six knowledge domains (**Figure 3a**). Both groups rated questions in the medical domain (treatment, health information) as most important, followed by the physical domain (symptoms, side effects). Caregivers rated approximately half of the items in the practical, psychological, and social domains as “very important”. Compared to caregivers, patients’ importance scores were higher in the psychological and social domains, and lower in the practical domain. Both groups rated spiritual needs as least important. In all domains, importance score differences between groups were not significant (t≤1.56, *p*≥0.12). Overall, the items rated as most important by both caregivers and patients belonged to the medical and physical domains, and included information about symptoms and side effects, treatment options, and medication (**Supplementary Table 2**).

**Figure 3.**
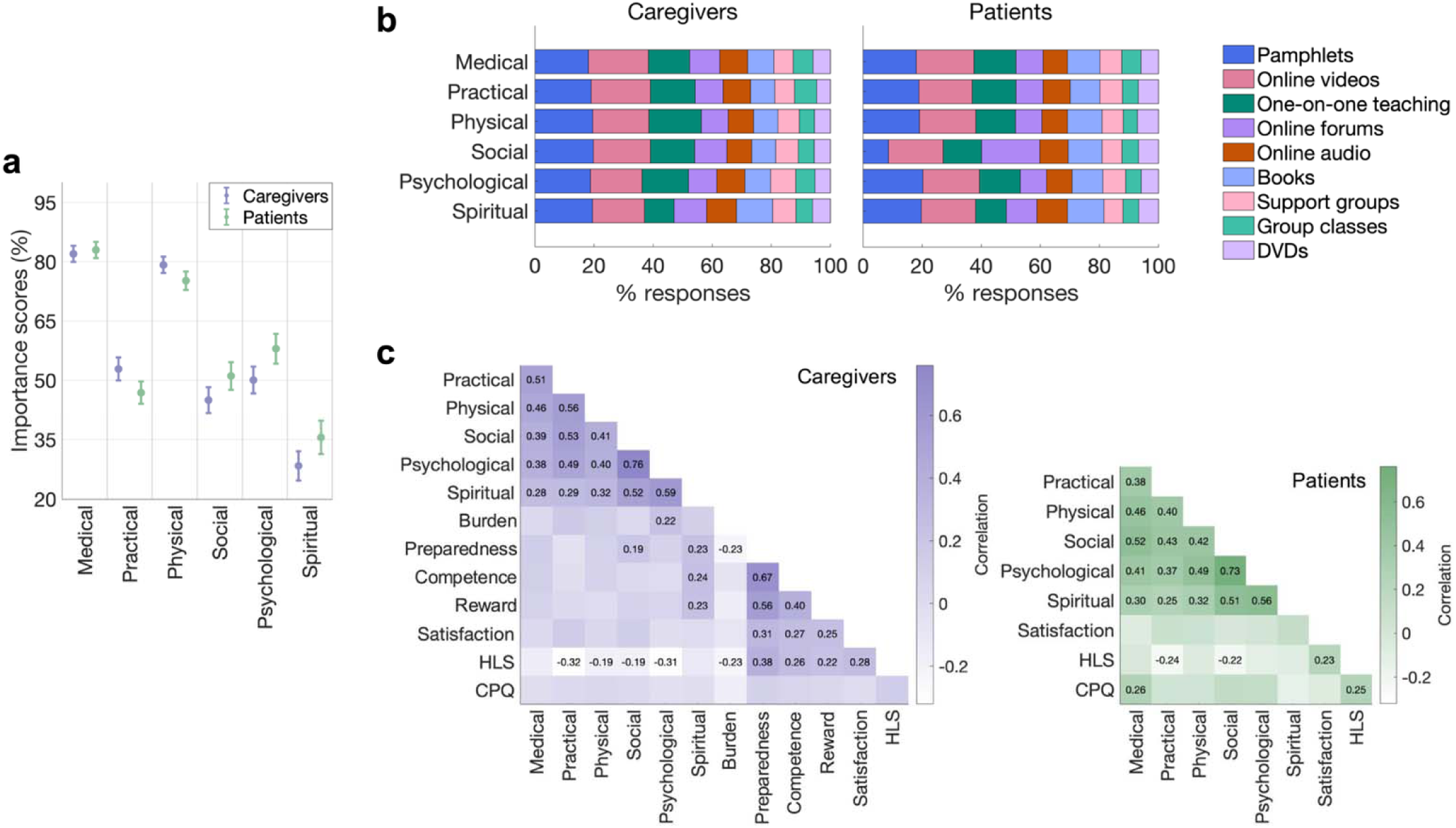
Caregiving-related informational needs. **a.** Importance scores (percentage of items rated as “very important”) in each domain by caregivers and patients. Markers show means; error bars are SEM. **b.** Preferred information delivery methods. Stacked bars show the percentage of responses indicating a preference for each method. Note that participants were able to choose multiple methods. **c.** Correlations between domain importance scores, caregiving scale scores (for caregivers), satisfaction with care, health literacy scores (HLS), and computer proficiency scores (CPQ). Only significant correlation values (*p*≤0.05) are displayed.

In both samples, pamphlets and online videos were the most popular methods of preferred information delivery (over 17% on average across domains), followed by one-on-one teaching, online forums, books and online audio materials such as podcasts (**Figure 3b**). Participant choices were consistent across domains, except for the social domain, where patients ranked online forums (19.7%) as their top choice.

### 3.9. Associations between informational needs and other variables

In both caregivers and patients, health literacy negatively correlated with informational needs (**Figure 3c**). In patients, health literacy also moderately correlated with computer proficiency. Satisfaction with care did not correlate with informational needs in either cohort. However, caregivers’ satisfaction with care and health literacy positively correlated with their preparedness, competence, and reward caregiving scores. Caregivers with higher health literacy also reported lower caregiving burden. Furthermore, in caregivers, spiritual domain scores correlated with caregiving preparedness, competence, and reward. Social and psychological domain scores correlated with caregiving preparedness and burden.

Using a stepwise regression procedure, we identified the optimal combination of variables to model informational needs in each domain (**Table 2**). For caregivers, the final models included health literacy scores across all domains. Their informational needs were also predicted by their preparedness for caregiving (medical, physical, social, and psychological domains), caregiving competence score (spiritual domain), English language fluency (practical, physical, and spiritual domains), and age (practical domain). Patients’ informational needs were best predicted by computational proficiency alongside the perceived time burden of caregiving (medical and social domains), gender (social domain), income (spiritual domain), age, and health literacy (practical domain).

**Table 2.** Multivariate regression results. Only significant predictors in the final models are shown. VIF: variance inflation factors.

| Caregivers |  |  |  |  |  | Patients |  |  |  |  |
| --- | --- | --- | --- | --- | --- | --- | --- | --- | --- | --- |
|  | Variable | Estimate | CI | P-value | VIF | Variable | Estimate | CI | P-value | VIF |
| Medical | PCS | 0.65 | 0.12,1.17 | 0.017 | 1.16 | CPQ | 0.81 | 0.2,1.42 | 0.01 | 1.01 |
|  | HLS | -1.67 | -3.03, -0.31 | 0.017 | 1.16 | Time spent caregiving | 8.75 | 0.32,17.17 | 0.042 | 1.01 |
| Practical | Age | -0.43 | -0.8, -0.07 | 0.021 | 1.03 | Age | -0.6 | -0.98, -0.22 | 0.003 | 1 |
|  | English fluency | -20.81 | -34.52, -7.1 | 0.003 | 1.01 | HLS | -1.52 | -2.87, -0.16 | 0.029 | 1 |
|  | Income | 25.35 | -45.23, -5.46 | 0.013 | 1.01 |  |  |  |  |  |
|  | HLS | -2.55 | -4.33, -0.77 | 0.005 | 1.01 |  |  |  |  |  |
| Physical | English fluency | -19.91 | -30.06, -9.76 | <0.001 | 1.04 |  |  |  |  |  |
|  | Main caregiver | -12.79 | -24.49, -1.09 | 0.032 | 1.01 |  |  |  |  |  |
|  | PCS | 0.71 | 0.2,1.22 | 0.007 | 1.22 |  |  |  |  |  |
|  | HLS | -2.11 | -3.4, -0.81 | 0.002 | 1.18 |  |  |  |  |  |
| Social | PCS | 1.48 | 0.67,2.28 | <0.001 | 1.16 | Gender | -14.57 | -28.31, -0.84 | 0.038 | 1.06 |
|  | HLS | -3.44 | -5.52, -1.37 | 0.001 | 1.16 | Time spent transporting | 40.76 | 13.2,68.32 | 0.004 | 1.06 |
|  |  |  |  |  |  | CPQ | 1.26 | 0.26,2.26 | 0.014 | 1.06 |
| Psych | Gender | 16.39 | 3.2,29.58 | 0.015 | 1 |  |  |  |  |  |
|  | PCS | 1.12 | 0.32,1.93 | 0.007 | 1.17 |  |  |  |  |  |
|  | HLS | -4.35 | -6.43, -2.27 | <0.001 | 1.17 |  |  |  |  |  |
| Spiritual | English fluency | -19 | -37.64, -0.37 | 0.046 | 1.05 | Income | 27.33 | 0.89,53.77 | 0.043 | 1.04 |
|  | COMP | 4.91 | 2.08,7.75 | <0.001 | 1.13 | CPQ | -1.4 | -2.77, -0.02 | 0.047 | 1.13 |
|  | HLS | -2.52 | -4.78, -0.25 | 0.03 | 1.08 |  |  |  |  |  |

### 3.11. Open-ended responses

Caregivers and patients responded to questions asking what information was needed the most (43 and 28 responses respectively) and provided additional comments (33 and 22 comments respectively). Thematically, responses reflected a strong need for information and support (**Supplementary Figure 3**), particularly in navigating and discovering resources: “*it would be helpful to have all the information accessible in one location*” (C30), “*it takes my daughter a lot of research to find answers*” (P7). Both caregivers and patients highlighted challenges in getting support, and expressed a need for medical and physical information, practical support (transportation and finances), as well as social and psychological support (“*it is very rare to be asked how I am doing”,* C27). The most common suggestions were to facilitate conversations between healthcare professionals and caregivers, and to make access to medical information easier for caregivers where consent is provided.

## 4. Discussion

This cross-sectional survey investigated the caregiving-related perceptions and informational needs of cancer caregivers and patients. We used validated scales to assess caregiver burden and evaluated caregivers’ and patients’ satisfaction with the medical care and caregiver support they had received. Participants reported their caregiving-related informational needs across six knowledge domains and stated their preferred methods of information delivery.

Results revealed a mismatch between caregiver and patient perceptions of caregiving burden and the impacts of cancer on caregivers, as well as between their levels of satisfaction with the medical care received by patients. Both caregivers and patients reported a strong need for information in the medical and physical domains and indicated a preference for pamphlets and online videos. Health literacy predicted the informational needs of caregivers and was negatively associated with caregiver burden. Caregivers’ satisfaction with care was associated with their caregiving preparedness, competence, and reward.

Our results highlight the interplay between health literacy, caregivers’ experiences, and their informational needs. The differences between caregivers’ and patients’ perceptions emphasize the need for tailored support for caregivers. Caregivers would benefit from more medical and physical information, as well as practical support and help navigating existing resources. These findings highlight the need for caregiver education, including the importance of interventions that aim to build health literacy.

### 4.1 Diverging perceptions of caregiving burden and medical care

Caregivers in our study reported moderate caregiving burden, with over 50% spending over 15 hours per week caregiving. While caregivers and patients had similar perceptions of the time spent by caregivers, patients were more likely to report that they needed no daily practical help. Previous work has suggested that patients may underestimate the level of support they need, perhaps out of a need to perceive themselves as independent^25^.

Compared to caregivers, patients also underestimated the impact of cancer on caregivers across all areas, from physical health to changes in values. This is in line with previous reports suggesting that patients may underestimate caregiver burden, including the emotional impacts and practical difficulties of caregiving^17,26^.

Both groups reported similar levels of satisfaction with the support caregivers had received, although fewer caregivers reported being involved or shown interest by healthcare professionals. Caregivers were also less satisfied than patients with the medical care received by patients. This is consistent with previous work^27^ and suggests that caregivers may feel unsupported by healthcare professionals^3,28^. The burden and stress of caregiving, including impacts on caregivers’ time, finances, and health, may contribute to their dissatisfaction with the healthcare experience. Furthermore, while patients often focus on the immediate impacts of their illness, caregivers may have higher expectations of healthcare services and be distressed by the patients’ experience^29^. Previous work has shown that caregivers overestimate patients’ symptom burden and pain intensity^25^, and that such differences in perception can lead to worse outcomes for both patients and caregivers^30^. It is thus critical that caregivers receive adequate support tailored to their needs, enabling them to feel more prepared and reducing the gap between patient and caregiver perceptions.

In line with previous reports, women reported higher levels of caregiver burden and more time spent caregiving^31^. In Canada, more than half of women provide care to family members (52% compared to 42% of men)^32^, and women caregivers report higher levels of depression^33^. To close this gap, support should be provided to help families distribute caregiving burden whenever possible, as well as protect women’s time and employment. Furthermore, other axes of marginalization that may increase caregiving burden should be considered to ensure equitable caregiver support^34^.

### 4.2 Caregivers need medical and physical information

Caregivers’ and patients’ assessments of the information needed by caregivers were remarkably similar across the six knowledge domains. Consistent with informational assessments conducted with cancer patients^11,12^, medical information was rated as most important by caregivers and patients, followed by physical information. Items highly ranked by both groups included medical and physical information about treatment options, symptoms, side effects, and medication, while items about the effects of cancer on appetite and fatigue were rated as important by caregivers. For patients, the psychological domain was the third most important, while caregivers placed more importance on the practical domain. These results reflect the varied and complex tasks involved in caregiving, which often lead caregivers to feel unprepared when supporting patients^2^. Open-ended responses also emphasized the need for medical and practical support, as well as the challenge of navigating an often-overwhelming number of resources and information.

Both groups rated spiritual-domain items as least important. However, spiritual domain scores correlated with caregiving preparedness, competence, and reward, in line with work linking spirituality and caregiving coping skills^35^.

### 4.3 Caregivers and patients prefer pamphlets and online videos

Both caregivers and patients expressed a preference for pamphlets and online videos, highlighting the importance of self-paced, asynchronous learning. The popularity of both online and physical media highlights the need for diverse information streams. Alongside digital materials, pamphlets maintain their value as tangible, accessible, and compact resources even in samples with high computer proficiency^37^. One-on-one teaching was also highly ranked by participants, in line with open-ended comments calling for more one-on-one discussions between caregivers and healthcare professionals.

### 4.4 Health literacy impacts caregiving perceptions and needs

Caregivers with higher health literacy reported fewer informational needs, lower caregiver burden, and higher caregiving preparedness, competence, and reward. These results highlight the role of health literacy in helping caregivers navigate information and feel prepared in their role. Alongside health literacy, English fluency and preparedness for caregiving also predicted informational needs. Previous work has found associations between health literacy and caregiver or patient outcomes^38^, and has shown that caregivers from culturally and linguistically diverse communities struggle to navigate health information^39^. Low health literacy may make it difficult for caregivers to understand the patient’s medical condition, treatment plan, or prognosis, as well as to seek and obtain the necessary information to effectively manage the patient’s care. This may increase caregiving burden and feelings of unpreparedness and overwhelm.

Strengthening health literacy should thus be a priority in efforts to support caregivers. Yet anticipating caregivers’ medical information needs requires careful consideration, and the context of life-threatening illness may make it difficult for caregivers to learn. Efforts to improve cancer health literacy among the public would mitigate this and help caregivers feel more prepared when taking on their new role. Given the prevalence of online misinformation and the information overload reported by cancer caregivers, health literacy interventions should also address digital health literacy, particularly among disadvantaged groups^40^.

### 4.5 Clinical implications

While programs for caregivers often focus on psychological support^5^, these results highlight a strong need for medical, physical, and practical information. With an increase in outpatient cancer care, caregivers perform more tasks that require medical and practical knowledge, such as administering medicine, helping manage pain, or deciding when to seek help^3^. The complexity of these tasks may increase caregiver burden and burnout, leading to the psychological impacts often reported in the literature^7,36^. To improve caregiver and patient outcomes, interventions should focus proactively on practical and medical support and information alongside psychological support.

### 4.6 Limitations

The surveys did not require or record the presence of caregiver and patient dyads, and thus we can only discuss differences in perceptions at the group level. Only English-speaking patients and caregivers were recruited; however, language-diverse populations may have very different experiences, and indeed our findings highlight a connection between English fluency and informational needs. Furthermore, while the present data reflect a diverse sample with a wide range of socio-demographic characteristics, cancer types and stages, and caregiving burdens, this may limit our ability to identify specific challenges facing different groups. However, the patient and caregiver samples were well-matched in terms of cancer type and stage and other patient characteristics, enabling comparisons between groups.

### 4.7 Conclusion

We evaluated the perceptions and informational needs of cancer caregivers, as well as patients’ perceptions about caregivers’ needs. While caregivers and patients agreed on informational needs, their perceptions of caregiving burden and medical care differed, highlighting potential gaps in caregiver support. Caregivers call for more medical and physical information and need better health literacy to face the challenges of caregiving. Together, our results highlight a strong need for tailored training and support for cancer caregivers and for the healthcare professionals interacting with them.

## Data Availability

The data that support the findings of this study are available on request from the corresponding author. The data are not publicly available due to privacy or ethical restrictions.

## Notes

### Competing Interest Statement

The authors have declared no competing interest.

### Funding Statement

This study was supported by the Princess Margaret Cancer Foundation.

### Author Declarations

The Institutional Research Ethics Board of the University Health Network gave ethical approval for this work (REB#: 09-0952).

