## Supplementary for "Strengthening support for cancer caregivers: an investigation of caregiving perceptions and informational needs"

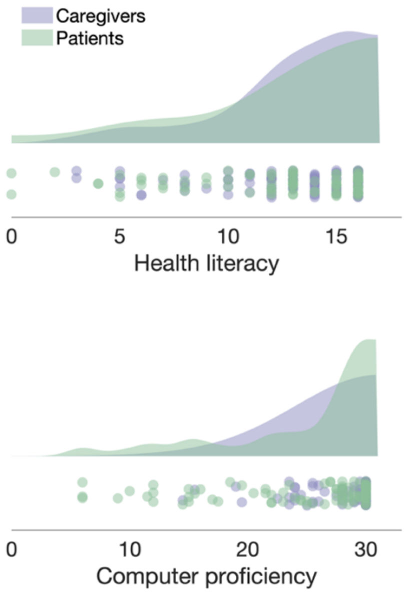


**Supplementary Figure 1**. Distribution of health literacy and computer proficiency scores among patients and caregivers. Individual data points are shown below the estimated distributions.

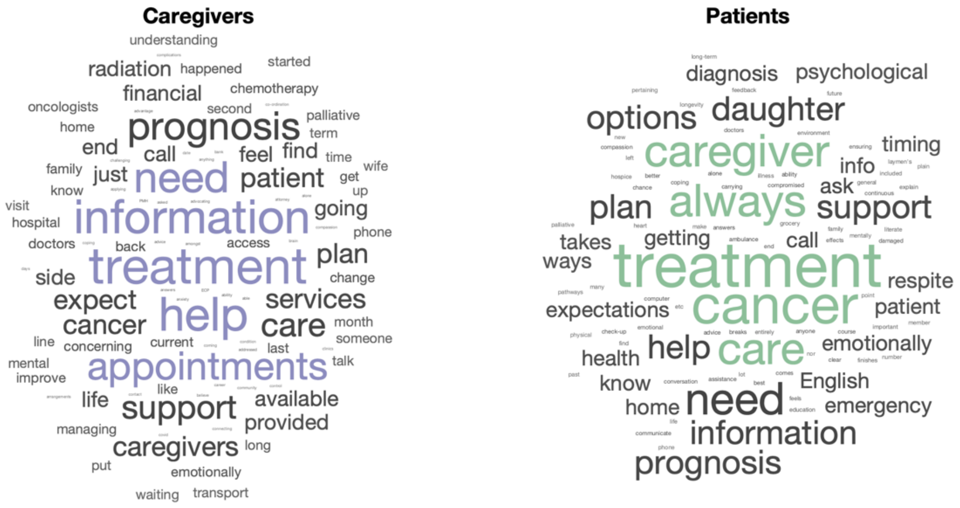


**Supplementary Figure 2**. Word clouds of the open-ended responses to the question, “What information or support do you need right now?” Words used more frequently are displayed in larger font size.

**Supplementary Table 1**. Perceptions of caregiver support received. The table displays the percentage of participants within each sample (caregivers and patients) that selected each answer option. HCP: healthcare professionals.

| **Question** | **Caregivers** | | | | | **Patients** | | | | |
|---|---|---|---|---|---|---|---|---|---|---|
| Have HCP shown an interest in how you have been feeling? (%) | Rarely/Never<br>32.7 | Only sometimes<br>21.2 | Most of the time<br>18.3 | Always/ Almost always<br>18.3 | Do not know<br>9.6 | Rarely/Never<br>26.7 | Only sometimes<br>14.8 | Most of the time<br>23.5 | Always/ Almost always<br>22.2 | Do not know<br>8.6 |
| Have HCP involved the caregiver in care in the way they wanted? (%) | Rarely/Never<br>12.5 | Only sometimes<br>15.4 | Most of the time<br>24 | Always/ Almost always<br>43.3 | Do not know<br>4.8 | Rarely/Never<br>4.9 | Only sometimes<br>9.9 | Most of the time<br>38.3 | Always/ Almost always<br>40.7 | Do not know<br>6.2 |
| Have HCP noticed that the caregiver wasn't doing well and offered help? (%) | Rarely/Never<br>30.8 | Only sometimes<br>7.7 | Most of the time<br>9.6 | Always/ Almost always<br>12.5 | Do not know<br>39.4 | Rarely/Never<br>24.1 | Only sometimes<br>7.6 | Most of the time<br>7.6 | Always/ Almost always<br>11.4 | Do not know<br>49.4 |
| Has enough time been spent giving caregivers information? (%) | No, not at all<br>17.1 | Yes, to a low degree<br>18.1 | Yes, to some degree<br>28.6 | Yes, to a high degree<br>23.8 | Do not know<br>12.4 | No, not at all<br>12.3 | Yes, to a low degree<br>12.3 | Yes, to some degree<br>28.4 | Yes, to a high degree<br>30.9 | Do not know<br>16 |
| Has information been provided in a sensitive way? (%) | No, not at all<br>4.8 | Yes, sometimes<br>24.8 | Yes, most of the time<br>60 | Do not know<br>10.5 | | No, not at all<br>7.4 | Yes, sometimes<br>19.8 | Yes, most of the time<br>64.2 | Do not know<br>8.6 | |
| How well has the hospital informed the caregiver? (%) | Very poorly<br>6.7 | Pretty poorly<br>13.3 | Pretty well<br>38.1 | Very well<br>28.6 | Do not know<br>13.3 | Very poorly<br>1.2 | Pretty poorly<br>6.2 | Pretty well<br>39.5 | Very well<br>44.4 | Do not know<br>8.6 |

**Supplementary Table 2**. Top-ranked items across domains. Shaded questions are highly ranked by one group and not the other (purple: caregivers; green: patients). #: the number of participants rating each item as "very important".

| Rank | Caregivers | | | Patients | | | Rank |
|---|---|---|---|---|---|---|---|
| | Question<br>How important is it for you to have information about… | Domain | # | Question<br>How important is it for your friend/family member to have information about… | Domain | # | |
| 1 | Different treatment options and their advantages (like success rates) and disadvantages (like side effects and risk of death) | Medical | 105 | What symptoms and side effects to watch out for and to report them to your friend/family member's health care team | Physical | 86 | 1 |
| 1 | What symptoms and side effects to watch out for and to report them to your friend/family member's health care team | Physical | 105 | The follow-up visits and different medical tests you will need after treatment | Medical | 85 | 2 |
| 2 | The follow-up visits and different medical tests your friend/family member will need after treatment | Medical | 104 | Different treatment options and their advantages (like success rates) and disadvantages (like side effects and risk of death)? | Medical | 84 | 3 |
| 3 | How medications (drugs) should be taken by your friend/family to decrease their risk (chance) of facing side effects | Physical | 102 | How you should take medications (drugs) to decrease your risk (chance) of facing side effects | Physical | 83 | 4 |
| 4 | How to manage the pain your friend/family may experience due to cancer | Physical | 99 | The health care workers involved in your care and how they can help (like a surgeon, radiation oncologist, dietitian, nurse, radiation therapist, speech language pathologist or social worker) | Medical | 81 | 5 |
| 4 | Changes to your friend/family member's appetite due to cancer and its treatment | Physical | 99 | General cancer information (like your cancer type) | Medical | 78 | 6 |

| | | | | | | | |
|---|---|---|---|---|---|---|---|
| 5 | How your friend/family member can manage cancer-related fatigue (a feeling of tiredness that does not go away with rest or sleep) due to cancer and its treatment | Physical | 97 | How often you should visit their doctor during treatment | Medical | 77 | 7 |
| 6 | How your friend/family member can manage nausea (feeling of having to throw-up) and vomiting (throwing up) due to cancer | Physical | 96 | How to manage the pain you may experience due to cancer | Physical | 77 | 7 |
| 7 | General cancer information (like your friend/family member's cancer type) | Medical | 94 | Your hidden or long-term side effects from treatment | Physical | 77 | 7 |
| 8 | How often your friend/family member should visit their doctor during treatment | Medical | 93 | How to manage your nausea (feeling of having to throw-up) and vomiting (throwing up) due to cancer | Physical | 76 | 8 |
| 8 | What to expect during end-of-life and the care your friend/family member will receive | Medical | 93 | | | | |
| 8 | The health care workers involved in your friend/family member's care and how they can help (like a surgeon, radiation oncologist, dietitian, nurse, radiation therapist, speech language pathologist or social worker) | Medical | 93 | | | | |
